# The Male Fertility Gene Atlas – A web tool for collecting and integrating data about epi-/genetic causes of male infertility

**DOI:** 10.1101/2020.02.10.20021790

**Authors:** H. Krenz, J. Gromoll, T. Darde, F. Chalmel, M. Dugas, F. Tüttelmann

## Abstract

Interconnecting results of previous OMICs studies is of major importance for identifying novel underlying causes of male infertility. To date, information can be accessed mainly through literature search engines and raw data repositories. However, both have limited capacity in identifying relevant publications based on aggregated research results e.g. genes mentioned in images and supplements. To address this gap, we present the Male Fertility Gene Atlas (MFGA), a web tool that enables standardised representation and search of aggregated result data of scientific publications. An advanced search function is provided for querying research results based on study conditions/phenotypes, meta information and genes returning the exact tables and figures from the publications fitting the search request as well as a list of most frequently investigated genes. As basic prerequisite, a flexible data model that can accommodate and structure a very broad range of meta information, data tables and images was designed and implemented for the system. The first version of the system is published at the URL https://mfga.uni-muenster.de and contains a set of 46 representative publications. Currently, study data for 28 different tissue types, 32 different cell types and 20 conditions is available. Also, ∼5,000 distinct genes have been found to be mentioned in at least ten of the publications. As a result, the MFGA is a valuable addition to available tools for research on the epi-/genetics of male infertility. The MFGA enables a more targeted search and interpretation of OMICs data on male infertility and germ cells in the context of relevant publications. Moreover, its capacity for aggregation allows for meta-analyses and data mining with the potential to reveal novel insights into male infertility based on available data.

## Introduction

Male infertility is a prevalent and highly heterogeneous disease for which the underlying causes can currently be identified for only about 30% of male partners in infertile couples (Tüttelmann *et al*., 2018). In the last years, a growing number of studies have been published on the genetics and epigenetics of male infertility using a broad range of genomic technologies. For an overview see Oud *et al*. (2019). Often, researchers give extensive insight into their findings by providing supplementary data or access to their raw data. Theoretically, this enables clinicians and other researchers to validate those findings or interpret their own results in a broader context; in practice, however, the findability and reusability of this vast set of information is limited. To date, there is no public resource for the field of male infertility that enables clinicians and researchers alike to easily access a comprehensive overview of recent findings, although tools for other diseases have long been established, such as, for example, the Gene ATLAS (Canela-Xandri *et al*., 2018) that was released in 2017. It provides information on genetic associations of 778 traits identified in genome-wide association studies (Canela-Xandri *et al*., 2018), male infertility is, unfortunately, not one of them.

Instead of being accessible through a specified tool, research on male infertility relies mainly on general search engines for scientific publications like PubMed or Google Scholar. These engines can be employed to search for information based on keywords of interest, e.g., gene names in combination with specific conditions. However, if the required information can only be found in a supplementary table or figure, these search engines are unable to mark the corresponding publication as relevant. Other sources of information are raw data repositories like Gene Expression Omnibus (GEO) which provides access to the raw data files of microarray and genomic data from many publications (Barrett *et al*., 2013), or Sequence Read Archive (SRA). This is a marked achievement for the scientific community, but such repositories do not allow users to find a data file based on a gene or variant of interest. For the most part, publications of interest have to be identified in advance, and comprehensible insight into the data can only be achieved by reanalysing it, using complex bioinformatics pipelines.

To address this need specifically for the reproductive sciences, Chalmel and colleagues established the ReproGenomics Viewer (RGV) in 2015 (Darde *et al*., 2019; Darde *et al*., 2015). It provides a valuable resource of manually-curated transcriptome and epigenome data sets processed with a standardised pipeline. RGV enables the visualisation of multiple data sets in an interactive online genomics viewer and, thus, allows for comparisons across publications, technologies and species (Darde *et al*., 2019). However, the RGV is not designed to show downstream analysis results such as differentially expressed genes or genomic variation. Also, it is not possible to identify relevant publications based on genes of interest. Other tools in this field are GermOnline (Lardenois *et al*., 2010) and SpermatogenesisOnline (Zhang *et al*., 2013). However, both have not been maintained in recent years.

In order to offer a public platform that provides access to a comprehensive overview of research results in the field of epi-/genetics of male infertility and germ cells and to bridge the gap between textual information in publications on the one hand and complex data sets on the other hand, we designed, developed, and now introduce the publicly available Male Fertility Gene Atlas (MFGA, https://mfga.uni-muenster.de). Its objective is to provide fast, simple and straightforward access to aggregated analysis results of relevant publications, namely by answering questions like “What is known about the gene *STAG3* in the context of male infertility?” or “Which genes have been identified to be associated with azoospermia?”. To this end, we created an advanced search interface as well as comprehensive overviews and visualisations of the publications and search results. A basic prerequisite was that we had to design and implement a data model that can accommodate and structure a very broad range of meta-information, data tables and images.

## Materials and methods

### Requirements engineering

The MFGA is designed to support researchers and clinicians in the field of male infertility in finding and evaluating the analysis results of relevant publications. To ensure that the MFGA offers a relevant scope and focusses on the central needs and interests of the targeted user group, an extensive requirements engineering process had to be employed. This process aimed at defining the most relevant parts of the system design; in case of the MFGA, this means answering the following five questions:

1. What kind of data should be available?
2. How should the database be maintained and kept up to date?
3. Who should be authorised to access the data on the website?
4. What interface should be provided to access the data?
5. How should the data be visualised?

Since user-centric design is proven to be a critical factor of success for software projects (Maguire and Bevan, 2002), the requirements were acquired in close cooperation with a large group of prospective users of the MFGA. These researchers and clinicians are part of the Münster-based clinical research unit (CRU326) (http://www.male-germ-cells.de) and provided input on a broad range of aspects of male infertility, germ cell development as well as sperm morphology, motility, and function. The development of the MFGA is based on the software development and requirements engineering paradigm Rapid Prototyping, which focusses on developing early testing versions of the product to enable informed user feedback and iterative improvements (Budde *et al*., 1992; Gordon and Bieman, 1995).

The requirements analysis was based on a set of 39 representative and relevant publications/data sets that the MFGA should be able to host, provided by members of the CRU326. The publications were highly heterogeneous and covered a broad range of methodologies, from GWA studies to targeted genotyping, microarray, bulk or single-cell RNA sequencing as well as methylation. The complete list is available in Suppl. Tab. S1 and has already been included into the MFGA database.

### Current state of requirements

The requirements regarding the kind of data that should be available in the MFGA can be summarised in two points: First, the MFGA should host data representing the analysis results of a comprehensive set of publications on male infertility. Generally speaking, the MFGA needs to be able to deal with publications of arbitrary form and content. To account for the extremely high degree of heterogeneity between publications, a very flexible, modular data model is required. However, to enable the MFGA’s central service of offering selective data retrieval operations, it is crucial to identify, standardise and tag recurrent relevant information items and to curate them manually for each publication. Whenever applicable, standardisation should be based on ontologies as provided by OBO Foundry (Smith *et al*., 2007). Second, the MFGA should provide complementary data from external databases to increase the comprehensibility of information, e.g., to explain the background of gene names and provide links to more details. However, the MFGA is not supposed to contain any raw data; instead the system should link to the corresponding repositories if provided by a publication. Also, the MFGA is not planned for processing raw data by itself; the system is only intended to show aggregated result data provided by publications.

The database of the MFGA should be fully public and accessible to all researchers and clinicians in the field of male infertility. Maintenance and curation of selected publications should be provided by the development team. In order to keep the database up to date in a structured way, a recurring process for preparing new relevant publications will be implemented (Fig. 1). It comprises three main steps: selection of relevant publications, manual curation according to the requirements of the MFGA database and data upload. Prospectively, it should be triggered once per month and combine the proposed publications of four different sources into one prioritised list. The four information sources are (1) the list of not yet processed publications from the previous month, (2) newly published manuscripts from the CRU326, (3) other relevant new publications identified by the MFGA team, and, as an additional unbiased source, (4) recent publications queried from PubMed (query: “(male OR men) AND (fertility OR infertility) AND (gene OR genetic) AND (“YYYY/MM/01”[Date - Publication] : “YEAR/MM/31”[Date - Publication])”). Publications will be curated and uploaded by priority. Eventually, users are supposed to be enabled to act as registered contributors and support selection, curation and upload of content. Thus, a user management and authentication system is required.

**Figure 1.**
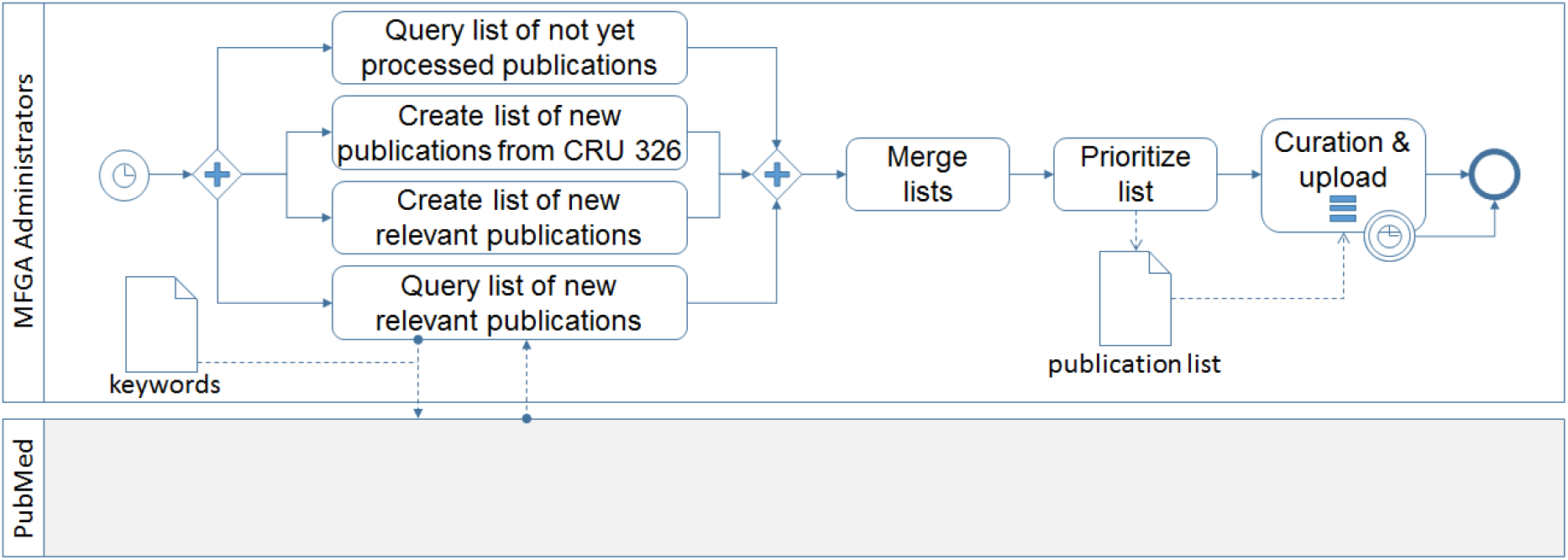
The process of identifying, selecting and prioritising novel publications depicted based on Business Process Model and Notation. (Chinosi and Trombetta, 2012). The process is triggered monthly. Potential publications are collected and merged into a list. This list is then prioritised by the MFGA team. Subsequently, curation and upload are performed in the determined order.

Regarding the interface for accessing the information, users should be able to view a complete list of publications and their key features to get a quick overview on the full content of the data base. Additionally, an advanced search function should provide identifying and showing subsets of relevant publications based on the occurrence of gene names or IDs in tables and figures as well as meta-information like data type, OMICS, processed tissues/cells and conditions/species. Detailed information and results of each individual publication should be displayed by comprehensible, standardised overviews, including images, tables and on-demand plots. As a special feature, the MFGA should display data from different publications in an aggregated way to allow for meta-analyses and integrated analyses. All pages should be enriched with appropriate additional information from external databases and with cross references, allowing for quick navigation through the atlas.

## Results

### IT architecture

The MFGA has been implemented as a modern *Java* web application that is securely hosted on a server at the University of Münster, Germany, and can be accessed freely via the URL https://mfga.uni-muenster.de. Its architecture (Suppl. Fig. S1) is based on the specific requirements, identified during requirements engineering and explained in detail in the supplement. For a good user experience, the graphical user interface utilises the *Bootstrap* library which enables a smooth adjustment of the application to different browsers and devices (Otto *et al*., 2020) and familiar design elements. On the server side, Spring Security framework is employed to provide secure authentication processes and manage data access privileges (Pivotal Software Inc., 2019).

### Data model

The specific relational data model of the MFGA as well as the preparation process for publication data are a direct result of the requirements analysis and were developed based on the initial set of relevant publications. They are the main prerequisite for enabling complex search queries, aggregation functions and meta analyses on the MFGA’s database. For representing the highly heterogeneous publications, a flexible and thus modular data model was designed (Suppl. Fig. S2). It is based on information items that were identified to be preserved over large subsets of publications on male infertility and standardised to seven classes of information items: *publication meta information, data set meta information, processed tissue type, processed cell type, cohort’s condition, image* and *table* (Tab. 1). For each publication, they can freely be combined in any quantity including zero. In order to prevent inconsistencies in the database caused by misspelling or synonymous terms, structured data entry with drop-down lists and ontologies is implemented. Currently, the MFGA employs BRENDA Tissue Ontology (BTO) (Gremse *et al*., 2011) for tissue type definition, Cell Ontology (CL) (Diehl *et al*., 2016) for cell types and Human Phenotype Ontology (HPO) (Köhler *et al*., 2019) for conditions. Further technical specification, especially representation and annotation of data tables and images, is explained in the extended methods (see supplement).

**Table 1.** Information classes and items available for each publication.

| Information Class | Information items<sup>1</sup> |  |
| --- | --- | --- |
| Publication meta information | title | publishing date |
|  | author | important genes* |
|  | citation | link to publication |
|  | abstract | link to repository* |
| Data set meta information | title* | OMICS type |
|  | technology* | reference genome* |
|  | data type | species |
| Processed tissue types | name | maturity* |
|  | description* | potential* |
|  | no of subjects* | species |
|  | no of probes per subject* | reference genome* |
| Processed cell types | name | maturity* |
|  | description* | potential* |
|  | no of subjects* | species |
|  | no of cells per subject* | reference genome* |
| Cohort's conditions | Human phenotype ontology* id (Köhler *et al.*, 2019) | name |
|  | size of cohort* | comment* |
| Images | title | description* |
|  | annotation of genes or relevant IDs | path |
| Tables | title | description* |
|  | annotation of standard columns (gene names, relevant IDs, loci, ...) | annotation of all additional columns as numeric or text |

<sup>1</sup>Optional items are marked with \*.

### Available content

Currently there are 46 publications available on the MFGA, and they comprise information on 101 data sets (Fig. 2), 28 different tissue types, 32 different cell types and 20 conditions. The majority of data sets contains data on the transcriptome (22 single-cell RNA sequencing, 12 bulk RNA sequencing & 3 microarray). The second largest group of data sets contains information on the genome / exome (13 targeted genotyping, 9 genome-wide association studies, 4 whole-genome sequencing & 4 sanger sequencing). Additionally, 6 data sets on whole-genome bisulfite sequencing and 2 on targeted deep bisulfite sequencing are available.

**Figure 2.**
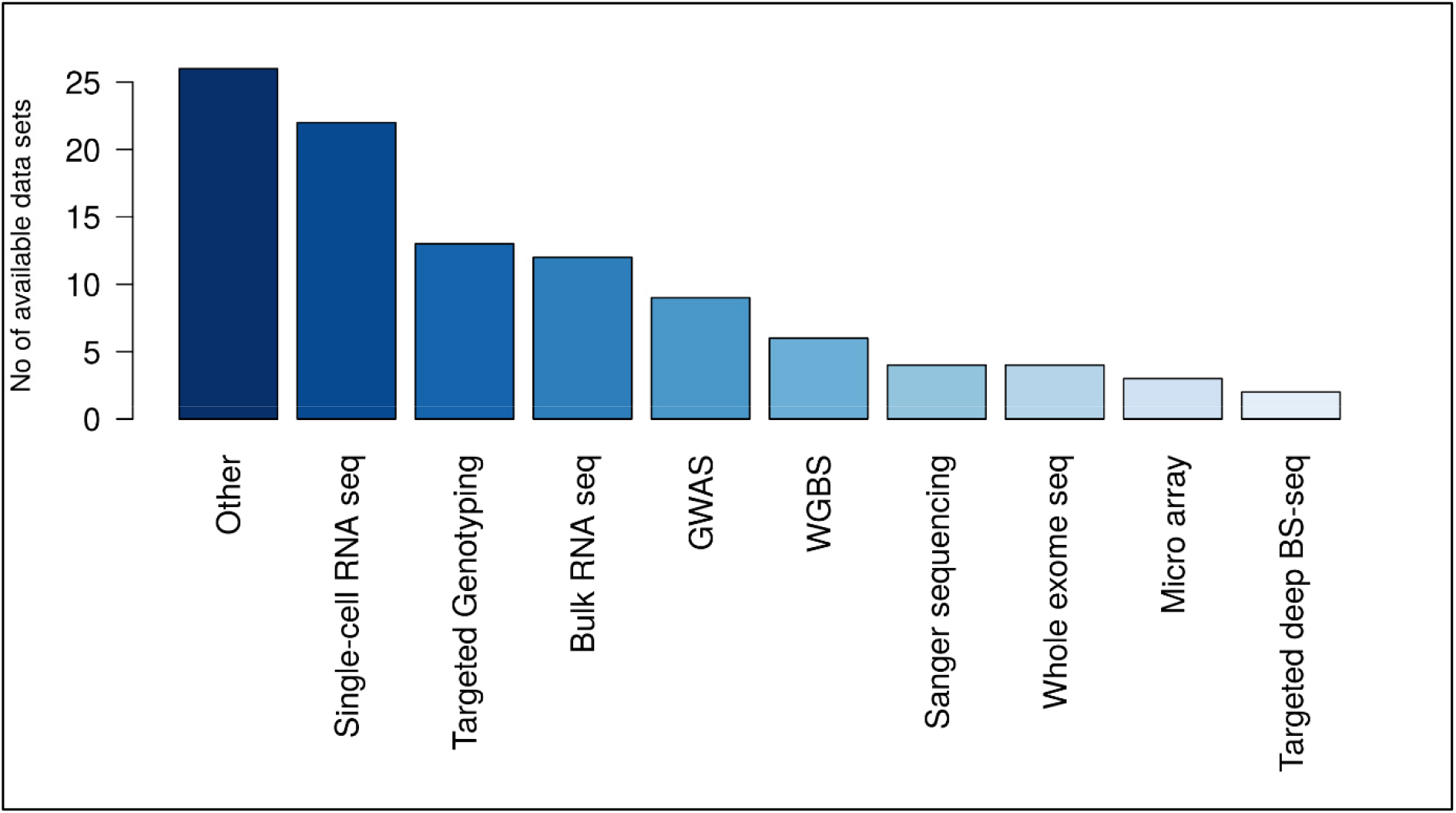
Data sets available on the MFGA grouped by technology. Currently 101 data sets corresponding to 46 publications are fully curated and publically accessible. Abbreviations: seq – sequencing, GWAS – genome-wide association study, WGBS – whole genome bisulfite sequencing, BS – bisulfite. Plot created with R (R Development Core Team, 2008).

The most frequent conditions in the data sets are variants of abnormal spermatogenesis, e.g. azoospermia and oligozoospermia. Testicular tissue has been processed by 32 proteome, methylome and transcriptome data sets. The predominantly represented cell types are embryonic stem cells, primordial germ cells, spermatogonia, spermatocytes, and spermatids. Also, ∼5,000 distinct genes have been found to be mentioned in at least ten of the publications, with the top genes being *TEX11, TEX14, TEX15, SYCP3, SMC1B, REC8* and *HORMAD1*. The full list of 46 publications can be accessed via the URL https://mfga.uni-muenster.de/publication.html.

### Functionality

The MFGA provides a web interface for fast, simple, and straightforward access to the results of publications on the epi-/genetics of male infertility and germ cells. For this purpose, an advanced search form is provided on the *Search* tab of the atlas (Suppl. Fig. S3), enabling the specification of search requests for relevant data sets. This function supports search terms from seven categories: OMICS, data type (e.g., single-cell RNA sequencing or targeted sequencing), condition, species, cell type, tissue type and gene/ID. For each of the first six categories, one can choose from a list of search terms equivalent to the information items stored in the database. Condition, cell type and tissue type are based on appropriate ontologies: HPO (Köhler *et al*., 2019), CL (Diehl *et al*., 2016) and BTO (Gremse *et al*., 2011). The number of available data sets is shown in brackets after each search term. Gene names and IDs can be specified in a text field as a comma-separated list. Also, there are three search modalities: (1) users can perform a broad search to identify all data sets that are annotated with at least one of the specified search terms, (2) users can perform a more targeted search for all data sets that are annotated with at least one of the specified search terms per category and (3) users can perform a very specific search for all data sets that are annotated with each of the search terms (default option). As an example, Fig. 3 shows an extract of the output of a simple search request for data sets in the MFGA database containing information on the gene *STAG3* (Suppl. Fig S2 for the full screenshot). This refers back to the introductory question: “What is known about the gene *STAG3* in the context of male infertility?”. For the following examples, the gene *STAG3*, which is located on chromosome 7, encodes a protein involved in meiosis and can be related to male infertility (van der Bijl *et al*., 2019), is employed to explain the general functionalities of the MFGA. A detailed *Walk Through* is provided on https://mfga.uni-muenster.de/walkThrough.html.

**Figure 3.**
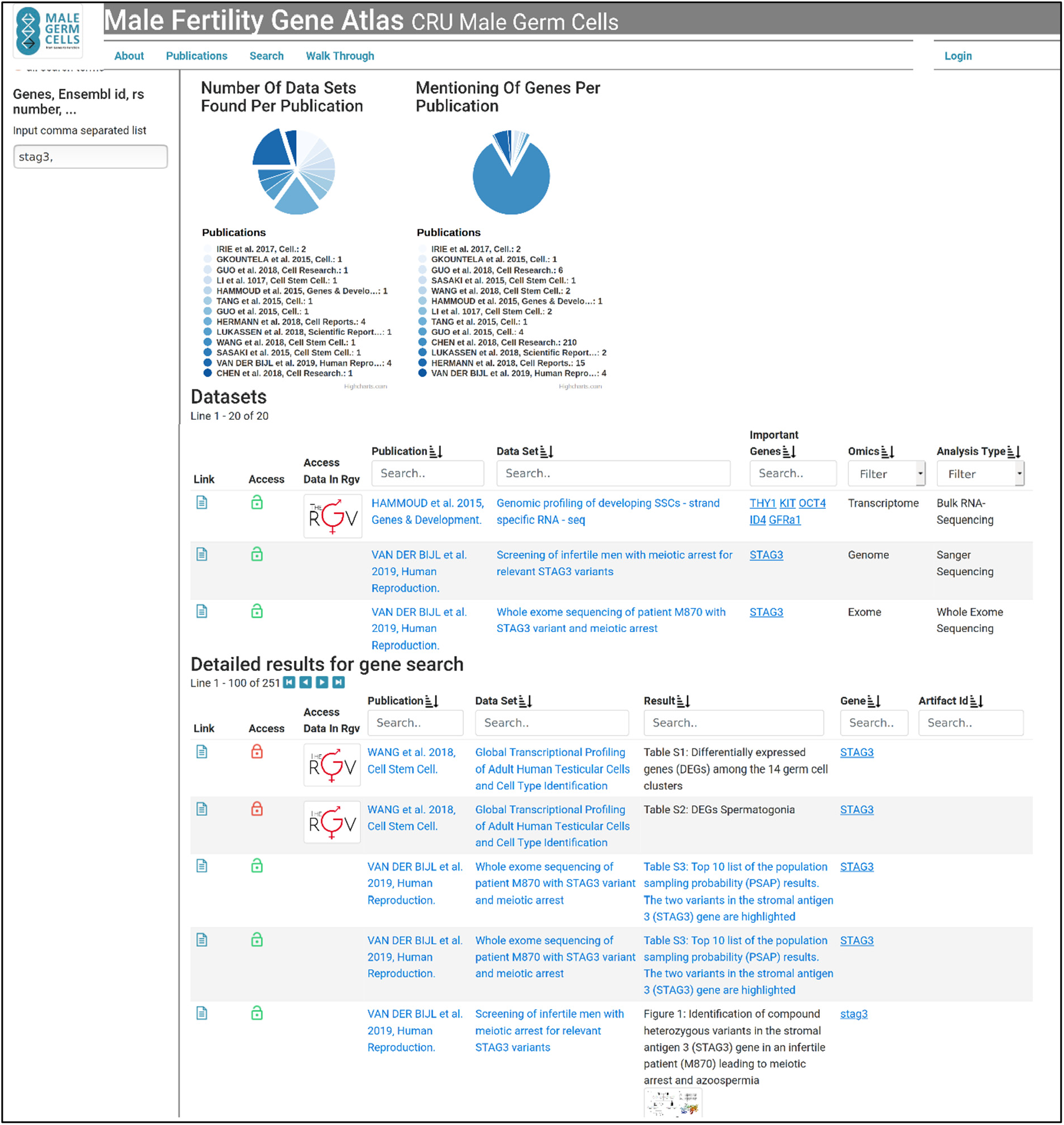
Example screenshot of the MFGA’s search functionality. (https://mfga.uni-muenster.de/search.html). It shows part of the result of the search for the gene *STAG3* in the MFGA database. The side bar on the left contains the search form. Here, *STAG3* is typed into the search field and search option ‘all search terms’ is checked. In the main window, plots show the number of returned data sets and the frequency of *STAG3* in data tables of the corresponding publications. Below, the returned data sets are presented in a table enriched with further meta-information, linking to the overview of data set and publication. The second table shows the specific data tables and images in which the gene was found. Here, tables are cropped and screenshot is compressed for better representability. See Suppl. Fig. S3 for original screenshot. Retrieved 10 January 2020.

Executing a search request on the MFGA results in a list of data sets matching the requirements, represented via charts and result tables (Fig. 3 and Suppl. Fig. S3). The left chart shows the number of returned data sets grouped by publication. Multiple data sets matching the search request in one publication can indicate that the authors provided further proof for their findings, such as e.g., van der Bijl *et al*. (2019), who screened two cohorts of infertile men. A textual summary of the data sets returned by the search request and some important meta-information is given by the *Data sets* table. All tables in the MFGA format are fully interactive such that the columns can be sorted, filtered and plotted online. Whenever the search contains genes or IDs, the right chart represents their frequencies in the different publications. This provides an initial estimation of relevance. Additionally, a second table lists their specific occurrences in data tables and figures. In the case when a search request is targeted at revealing relevant genes, such as, e.g., in the introductory question: “Which genes have been identified to be associated with azoospermia?” (Suppl. Fig. S4), the right chart and second table present the genes that occur most frequently in data tables of the returned data sets.

The search results returned by the MFGA are designed for quick navigation and information access. Therefore, many cross references are provided: Table entries directly link to overview pages of the corresponding publication and its data sets. Data tables and images are linked in the MFGA as well. However, this functionality is restricted to publications that are published under an open access license. As an example, Fig. 3 shows that the publication van der Bijl *et al*. (2019) mentions *STAG3*. Clicking on the data set title leads to the overview page of that publication (Suppl. Fig. S5). Also, data tables containing the searched gene can directly be accessed in MFGA by a single click, e.g., supplementary table 3 of van der Bijl *et al*. (2019) which is shown partly in Fig. 4 (full version in Suppl. Fig. S6). For a deeper analysis of the underlying read data of individual data sets, the MFGA provides a link to the RGV, whenever a data set is available in both tools, such as, e.g., Hammoud *et al*. (2015) in Fig. 3.

**Figure 4.**
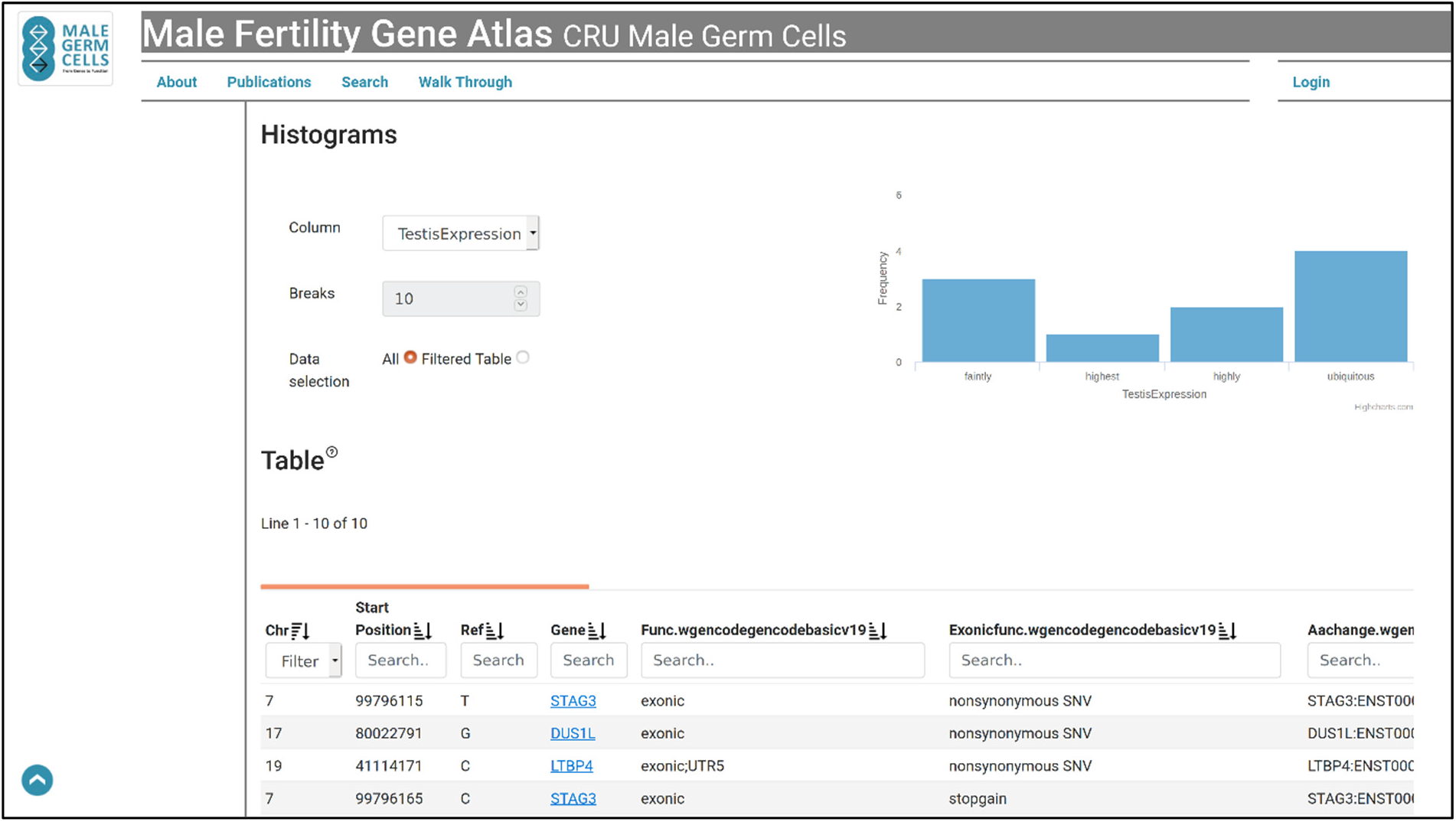
Example screenshot of the MFGA’s functionality to represent data from publications. (https://mfga.uni-muenster.de/publication/results?ssDetailsId=3968811). It shows part of supplementary table 3 of van der Bijl *et al*. (2019) in MFGA format. The table is represented in an interactive way and can directly be filtered and sorted. Additionally, histograms can be plotted for the table columns, here, e.g., based on column “Testis Expression”. For the full screenshot see Suppl. Fig. S6. Retrieved 10 January 2020.

Data from external public databases has been integrated in order to enrich information on the publications shown in the MFGA. Throughout the application, gene names can be selected in table cells and plots to open up an overlay with further explanations (for an example, see Fig. 5). The overlay bundles textual information from RefSeq (O’Leary *et al*., 2016) and HGNC (Yates *et al*., 2017; HGNC Database, 2018). Additionally, a link to the corresponding GeneCards page (Stelzer *et al*., 2016) is provided, as well as a shortcut to the MFGA search for that gene. Data from the GTEx project (Lonsdale *et al*., 2013; The Broad Institute of MIT and Harvard, 2019) is employed to show the gene’s expression in different tissues scaled by the largest median. The expression of the gene in testis tissue is highlighted in red.

**Figure 5.**
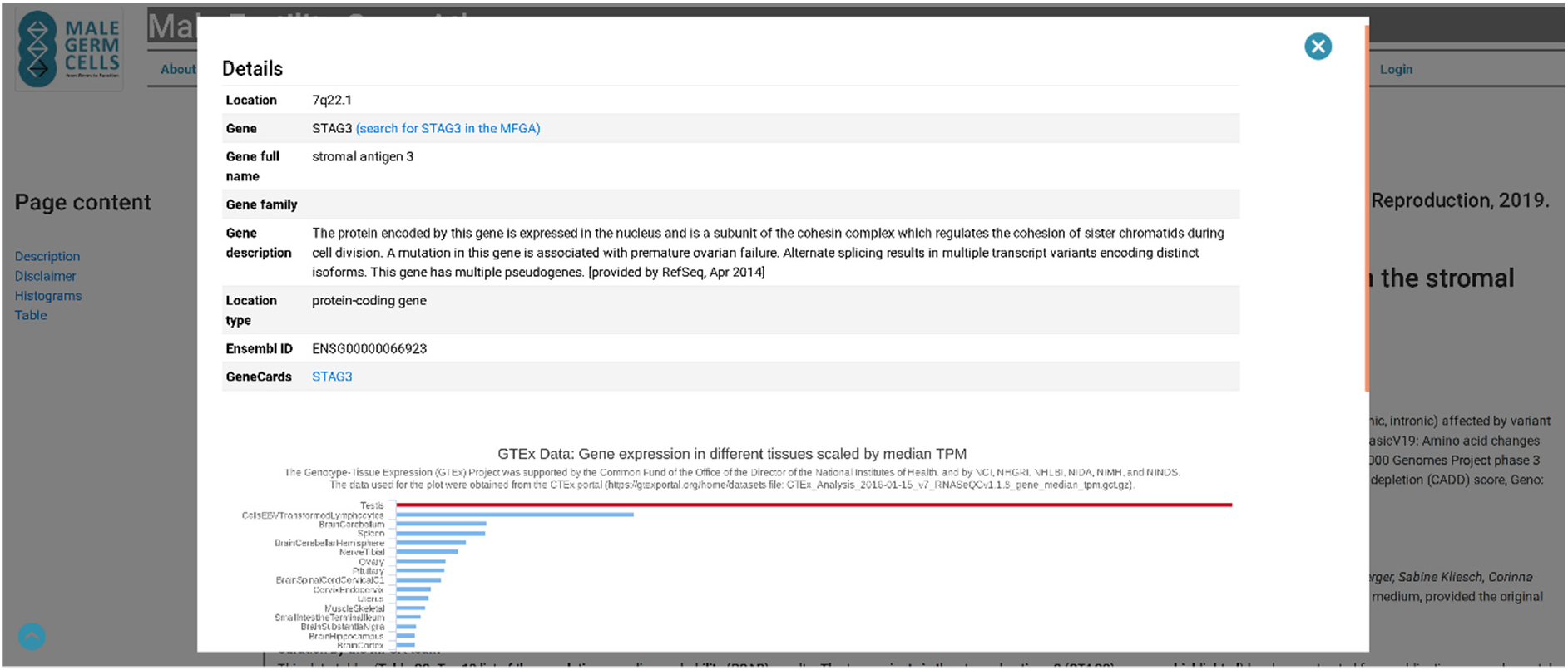
Screenshot of the MFGA showing the overlay for additional information on genes. (https://mfga.uni-muenster.de/publication/results?ssDetailsId=3968811). Here, as an example, the overlay for the gene STAG3 is shown. The textual information originates from RefSeq (O’Leary *et al*., 2016) and HGNC (Yates *et al*., 2017). Additionally, gene expression in different tissues is shown based on the data from the GTEx project (Lonsdale *et al*., 2013). Links for searching the gene in the MFGA database and opening the corresponding GeneCards entry (Stelzer *et al*., 2016) are provided. Retrieved 10 January 2020.

## Discussion

The MFGA provides a public platform to support researchers and clinicians in obtaining an overview of the recent findings in the field of male infertility and germ cells. The web-based tool includes recent libraries and methods for an improved user experience and straightforward usability. In order to enable an advanced search for relevant publications, a relational data model has been developed and implemented for knowledge representation. It records the recurrent information items in a structured and consistent way and can accommodate arbitrary publications from the field of male infertility, regardless of the kind of data analysis and technology. The search form enables a broad range of simple to very complex search queries such as “What is known about the gene *STAG3* in the context of male infertility?”, “Which genes have been identified to be associated with azoospermia?” or “Which genes have been found to be expressed in Sertoli cells of human testis tissue using single cell RNA sequencing?” (Suppl. Fig. S7).

The main purpose of the MFGA is to enable researchers and clinicians to consider their own research results in the context of other relevant literature. To this end, the system enables a highly selective search for studies with comparable conditions and parameters and offers the means to quickly review their main analysis results. Additionally, searches for genes can be performed in order to identify data sets containing the corresponding genes in their analysis results. This functionality supports the validation of candidate genes. Further, supplementary information from various sources is embedded into the MFGA, e.g., RefSeq (O’Leary *et al*., 2016), HGNC (Yates *et al*., 2017; HGNC Database, 2018) and GTEx project (Lonsdale *et al*., 2013; The Broad Institute of MIT and Harvard, 2019).

Compared to general search engines like Google Scholar and PubMed, with millions of records, the MFGA will always return smaller numbers of search results. However, in the domain of male infertility, the relevance of the individual results in relation to the corresponding search terms used in the MFGA is expected to be greater than when using those same search terms in a large search engine. Search engines for literature are usually restricted to mining textual information of publications and cannot consider information that is presented in tables, images, or supplementary material. The MFGA, however, is specifically designed for that task, which is enabled by its key features: A standardised data representation and disease-specific manual curation.

Another problem occurs when relevant data sets are searched in raw data repositories like GEO or processed data resources like RGV (Darde *et al*., 2015; Darde *et al*., 2019). While the information that these tools are able to provide is much more detailed than overviews in the MFGA, they cannot be used to identify a data set based on, for example, a list of genes that are supposed to be differentially expressed, since raw data does usually not include that kind of annotation. The MFGA facilitates searching through aggregated result data and, thus, identifying relevant data sets. Once identified, such data sets might then be further investigated in the RGV (Darde *et al*., 2015; Darde *et al*., 2019) or by processing GEO data with bioinformatics tools. Since RGV (Darde *et al*., 2015; Darde *et al*., 2019) and MFGA complement each other in their functionality, the MFGA links to RGV whenever a data set is present in both tools; further, an even closer integration is planned.

Regarding tools that might appear similar to the MFGA, Zhang *et al*. (2013) provide a tool termed SpermatogenesisOnline for searching individual genes in the context of spermatogenesis based on a set of publications from 2012 and earlier. However, it remains unclear which publications were included and whether or not they have been updated since then. Another tool from Lardenois *et al*. (2010), GermOnline, provides functional information about individual genes and access to transcriptomics data from microarrays on germline development from 2008. Both tools have, to our knowledge, not been updated and are, thus, not very useful anymore.

There are some limitations to the approach of the MFGA. Since analysis results are not reproduced using in-house pipelines, the MFGA is restricted to the analysis results authors are presenting in their publications. In the case where a publication provides, e.g., only significant SNPs from a GWA study, the MFGA, too, reports only these. The only meta-analyses enabled are those that use the aggregation of information in the MFGA, e.g., the top score of gene appearances, to approximate gene importance; the information cannot indicate correlation or even causality. Thus, the MFGA focusses on proposing plausible research hypotheses. Finally, full functionality of the MFGA can only be provided for publications under an open access license. Publications that allow no or restricted reuse are only partly integrated.

Prospectively, the MFGA will be updated and enlarged based on the proposed content management process (Fig. 1). In this context, a web form will be implemented to enable users to propose publications that are, from their point of view, still missing. Additionally, a closer integration of RGV and MFGA is planned, e.g. by coupling the publication submission forms, as well as further tools for meta-analyses.

In conclusion, the MFGA is a valuable addition to available tools for research on the epi-/genetics of male infertility. It helps fill the gap between pure literature searches as provided by PubMed and raw data repositories like GEO or SRA. The MFGA enables a more targeted search and interpretation of OMICS data on male infertility and germ cells in the context of relevant publications. Moreover, its capacity for aggregation allows for meta-analyses and data mining with the potential to reveal novel insights into male infertility based on available data. Ultimately, by combining RGV and MFGA with AI methods, we aim to develop a powerful gene prioritisation system dedicated to male infertility similar to the GPSy tool (Britto *et al*., 2012).

## Data Availability

The Male Fertility Gene Atlas is publicly available.

https://mfga.uni-muenster.de/

## Acknowledgements

We are grateful to all members of the Clinical Research Unit (CRU) ‘Male Germ Cells’ who contributed through either identifying relevant publications, supporting publication curation or providing feedback during the development of the MFGA: Michael Storck, Marius Wöste, Nina Neuhaus, Sven Berres, Sandra Laurentino, Lina Franziska Lanuza Pérez, Corinna Friedrich, Maria Schubert, Nikithomas Loges, Isabella Aprea, Jana Emich, Eva Maria Mall, Johanna Raidt, Nadja Rotte, Alexander Busch, Sara Kim Plutta, Jascha Henseler, Anna Natrup. We thank Dr. Celeste Brennecka for language editing of the manuscript.

## Authors’ roles

H.K. designed and developed the MFGA data model and system architecture, implemented the tool and drafted the manuscript. J.G. provided critical input during the development of the MFGA and first drafts of the manuscript. T.D. and F.C. contributed to integrating the mutual links between RGV and MFGA. M.D. and F.T. contributed to the system design and supervised the whole study. All authors critically revised the manuscript and approved the final version.

